# A Rational-Choice Model of Covid-19 Transmission with Endogenous Quarantining and Two-sided Prevention

**DOI:** 10.1101/2020.08.27.20183350

**Authors:** Joydeep Bhattacharya, Shankha Chakraborty, Xiumei Yu

## Abstract

This paper offers a parsimonious, rational-choice model to study the effect of pre-existing inequalities on the transmission of COVID-19. Agents decide whether to “go out” (or self-quarantine) and, if so, whether to wear protection such as masks. Three elements distinguish the model from existing work. First, non-symptomatic agents do not know if they are infected. Second, some of these agents unknowingly transmit infections. Third, we permit two-sided prevention via the use of non-pharmaceutical interventions: the probability of a person catching the virus from another depends on protection choices made by each. We find that a mean-preserving increase in pre-existing income inequality unambiguously increases the equilibrium proportion of unprotected, socializing agents and may increase or decrease the proportion who self-quarantine. Strikingly, while higher pre-COVID inequality may or may not raise the overall risk of infection, it increases the risk of disease in social interactions.

## 1 Introduction

Since the COVID-19 pandemic started in central China in late 2019, the disease has infected entire continents, ravaged the lives of millions and destroyed economies, lives, and livelihoods. Short of a safe and effective vaccine, no end appears in sight. The primary transmission route for COVID is believed to be human-to-human via respiratory droplets, small droplets that are ejected when speaking, coughing, or sneezing or via direct contacts. The most common symptoms of patients infected with the virus include a lower respiratory tract infection with fever, dry cough, and dyspnea. Some 40% of people, even when they are infected, show no symptoms and can unknowingly spread the infection to others.^1^

This raises an interesting problem: When X goes out to meet Y, neither Xnor Y knows whether they are infected and whether they can infect each other. Faced with infection risk, people, naturally, seek ways to avoid catching the virus. Xmay choose not to go out to visit with Y, or she may but wear a mask. What do X and Y decide to do? How do these choices relate to their affluence, infection risks, and costs and benefits? How does the income distribution of the economy in which X and Y live affect the infection risks these people face?

This paper is aimed at offering a minimalist, analytical framework in which to pose these questions. It permits each agent, X and Y, to rationally choose whether to “go out” and whether to engage in any preventive protection. Two elements sets it apart from existing work. First, the model explicitly allows for some agents to unknowingly transmit infection without being symptomatic. And second, it considers two-sided prevention using non-pharmaceutical interventions: when X and Y meet, the probability of X catching the virus from Y depends not just on what X does to protect herself but also on what Y does.

Most work on infectious-disease transmission starts with some version of the Kermack and McKendrick (1927) Susceptible-Infected-Recovered (SIR) model. That model assumes that the population in which a pathogenic agent is active comprises three subgroups: healthy individuals who are susceptible (S) to infection, infected individuals (I) who can infect the healthy ones, and individuals who are removed (R) from the infection cycle either due to death, immunity, or recovery. An infected individual randomly interacts with other susceptible agents and transmits the disease at a specific rate *β*; an infected individual also transits from the infection cycle, say via death or acquired immunity, at a particular rate, *γ*. These rates are taken parametrically both by the agents and the modeler, are calibrated using past data, and fed into the model. The resulting model is a system of ordinary differential equations, typically nonlinear due to the interaction between the infected and susceptible. Often, numerical experiments are run to study the likely progression of the disease (see Atkeson, 2020 or Acemoglu et al., 2020) by specifying a time-varying path for the basic reproduction number, *R* = *β*/*γ*.

While mathematical models are key to predicting the evolution and the aggregate impact of infectious diseases, their value depends crucially on how well people’s behavior is specified (Chen, 2013). Some previous work by economists (Kremer, 1996; and later Boucekkine and Laffargue, 2010, and Chakraborty et al., 2010 for example) start with a version of the SIR model and augment it to allow people to make choices about prevention.^2^ These choices, in turn, affect the aggregate infection rate, producing a system of simultaneous feedback. Choices are influenced not only by budgetary limits and/or preferences but also by informational constraints: whether the individual knows she is infected or infectious, and whether the people with whom she socializes are themselves contagious (Gersovitz and Hammer, 2003). In short, *β* and *γ* are endogenously determined.

We split ways from most of the existing economic-epidemiology literature by modeling how people, uninformed about their COVID-status, can spread the infection to others.^3^ In our model, a fraction of the population shows symptoms, gets enfeebled, and enters strict quarantine as symptomatic and infected (SI). The rest are either asymptomatic and infected (AI) or asymptomatic and uninfected (healthy, AH). Importantly, the non-SI do not know which group they are in. This is vital because the isolation of infected individuals is key to controlling transmission, and the decision to isolate often depends on observable symptoms (fever, cough, temperature). As Moghadas *et al*. (2020) point out, the effectiveness of such symptom-based interventions depends critically on the fraction of asymptomatic infections. The generally accepted range of estimates for asymptomatic COVID transmission is between 17.9% to 30.8% of all infections.

There is another critical point of departure for us. Almost the entire economic-epidemiology literature, erstwhile targeted to study HIV infections, focus on one-sided prevention. As noted above, a significant route of transmission of COVID is via small droplets ejected when speaking, coughing, or sneezing. In our setup, a person from the non-SI group may invest in personal protective equipment (PPE), such as masks or face covers, to protect herself from infectious droplets ejected by others she meets. Such PPE also offers source control, blocking droplets ejected by the wearer. However, serious concerns about the acceptability and tolerability of such non-pharmaceutical interventions linger; we know that because not everyone who should be wearing a mask does so, at least not diligently. For us, the upshot is PPE offers two-sided protection, but not everyone likes using them; our model allows for choices to be made in this regard. Interestingly, source control is more critical because if people wear masks to dim their chance of innocently infecting others, everyone ends up being more protected.

The focus in our model, unlike most epidemiological ones, is on intra-period decision-making. By design, all the action happens within a single period. An SI person here does not recover or reinfect or achieve immunity or die. Additionally, the only health-status transition we allow is for an AH to become infected. Doubtless, our focus on a one-period environment rules out many vital, inter-temporal concerns relating to the incubation period of the virus or the ramp-up of testing or the slow evolution of herd immunity. Even with that caveat, a lot is going on.

Agents, in the model, are heterogeneous in their endowment (income), *w*. As the period starts, people know if they are in the SI pool or not. What they do not know is whether they are in the AI or the AH pools. Since these decisions are taken *ex ante*, any non-SI agent attaches a probability, *α*, that they are in the AI group and uses that probability to compute the expected utility from a particular choice. Such people face two extensive-margin decisions: the decision to socialize (“go out”) and the decision to engage in prevention. Going out and meeting someone brings joy but may raise the infection risk.^4^ Similarly, protection activities, done diligently, entail goods and utility losses but reduce the chance of infection. To keep the analytics tractable, we make several non-critical, simplifying assumptions such as linear preferences, uniform income distribution, and so on.

In the standard SIR model, two assumptions are made. First, everyone knows their infection status. Second, at the start of the period, the entire pool of infected (*I*) people is assumed to mix randomly with the whole pool of susceptibles (*S*); each *S* meets every *I* and vice-versa, so there are *S* × *I* meetings. Afterward, a fraction *β* of the *S* has left their pool and joined the *I* pool. (Before the next period begins, some from the infected pool recover and may acquire immunity (or not) or perish. The membership in the various pools, at the start of the following period, is different from what it was when the previous period started.) In sharp contrast, we posit that entire pools do not mix: two people meet, bilaterally and only once, and neither is aware *ex ante* of the other person’s infection status or protection choice. The probability X encounters Y depends on the choice of Y to go out. The probability that Y is infected depends, additionally, on α. We introduce two-sided prevention by specifying that the likelihood of X (a member of the AH) getting sick from a meeting with Y depends multiplicatively on the person-specific, protection choices of both X and Y.

Agents under rational expectations take the masses of the three groups, SI, AH, and AI, as given. An individual determines her infection risk, and in turn, her going-out and protection choices, based both on these masses and her position on the endowment distribution. One compelling case has relatively poor people socializing without protection, the middle-income people socializing with protection, and the relatively affluent not socializing at all. Another case has the poor socializing without protection and the rest under self-quarantine. The big plus of our minimalist framework is that it allows precise, analytical determination of each of these entities.

Several equilibrium possibilities arise. Some equilibria represent extremes - all AI or AH go out or self-quarantine, and everyone socializing eschews/adopts PPE. Depending on deep parameters, more reasonable looking “interior” equilibria, are also possible. Central to our analysis is the following trade-off: All else same, less (more) use of PPE will raise (reduce) infection risk but may keep more (less) people home. In equilibrium, the overall effect will depend on the size of the groups which depends on who protects and who steps out. Our flagship result relates epidemiology with the income distribution. We find a mean-preserving increase in the spread of the distribution of income unambiguously increases the equilibrium proportion of socializing agents who are unprotected and may increase or decrease the percentage of agents who self-quarantine. Strikingly, while higher pre-COVID inequality may or may not raise the overall risk of infection is ambiguous, it definitely increases the risk of disease in social interactions.

A few recent papers are closely related to ours. Chang and Velasco (2020) propose a similarly minimalist model of economic choices during a disease outbreak. In a two-period model, agents who have been tested and marked healthy choose whether to go out to work (in which case, they may get infected) or to stay at home, both today and in the future. The decentralized equilibrium is inefficient due to an externality: when making these choices, people do not take into account the impact of their choice on the spread of the disease.

Dasgupta and Murali (2020) study the effect of COVID spread on future inequality in a SIR setup. In their setup, high and low-skill workers may choose to work onsite or remotely. Still, production activity in the high-skill occupations is such that the high-skilled find it easier to select the remote (hence, lower infection) option. Blundell *et al*. (2020) study pre-COVID inequality in the U.K. and document how the ability to continue working safely and through lockdowns is distributed very unevenly by gender, ethnicity, education, and earnings. Similarly, recent English data reveals that the most economically deprived areas have twice the rate of deaths involving COVID than the most affluent (Palmer, 2020). Schmitt-Grohe *et al*. (2020), however, find that when it comes to getting tested, income disparities did not matter for residents of New York City.

We develop the model in section 2 and study prevention decisions in section 3. Section 4 constructs the equilibrium implied by those decisions. Section 5 delves into the model’s implications for disease transmission and inequality. Section 6 concludes. Proofs are relegated to the appendix.

## 2 The Model

We study the decisions of one period-lived atomistic agents *i* ∊ [0,1] of mass *N* =1 and a government. An agent is endowed with income, *w^i^* drawn from a 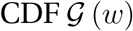 with finite support 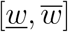. “High-*w*” agents are called “rich”, and low-w agents are “poor”.

At the start of a period, a fraction *h* of the population is asymptomatic: they do not feel sick, nor do they show signs of being sick. A fraction *α* of them, however, is infected with the virus, and may infect others; the rest, the truly “healthy” do not carry the disease. The fraction, 1 – *h*, is infected and symptomatic. In short, the entire population is split into three pools:

- *Asymptomatic-healthy* (AH) who are (1 – *α*) *h* fraction of the population,
- *Asymptomatic-infected* (AI) who are *αh* of the population, and
- *Symptomatic-infected* (SI), (1 – *h*) fraction of the population.

Transitions between these pools are restricted by assumption. We assume only the AH can, at a cost, increase the odds of preserving their health status and remaining in the AH pool; if they fail, they become infected. For tractability, this is the *only* state transition allowed.^5^

As the AI are unaware of their health status, they behave much like the AH, except they are infectious. However, both *α* and *h* are common knowledge. In what follows, we remove the current SI from consideration by assuming they are too debilitated to venture out, or because their apparent symptoms mandate government-enforced quarantine.^6^

### 2.1 Protection and social outings: costs and benefits

Healthy agents may preserve their health status by engaging in costly, preventive activities. These activities can take two forms: highly effective *self-quarantine* and other less effective forms that we will call *protection*. Protection includes a range of safety measures such as regular, diligent use of PPE – masks, face shields, gloves - hand sanitizers, and physical distancing while out and about. To keep things simple, we model the protection decision, e, as a discrete choice, *e ∊* {0,1} and *e* = 1 entails a goods cost *λ* > 0 reflecting the monetary cost of self-prevention. There is an additional, disutility cost, *k* > 0. We take the view that all prevention is self-protection, aimed mainly at reducing one’s *own* exposure to the disease. As noted earlier, use of PPE also offers source control, thwarting infectious droplets ejected by the wearer. We assume agents are self-interested and ignores this last bit.

The choice to go out is denoted by *μ* where *μ* ∊ {0,1}; *μ* = 0 means the agent is under self-quarantine (“public avoidance”), and *μ* =1 means the person goes out, once, and randomly socializes with one other person.^7^ Self-quarantine implies a fixed loss of κ units of income due to reduced access to production opportunities. Clearly, the cost of self-quarantine is disproportionately high for the poor.^8^ Going out, on the other hand, generates a fixed utility gain, *x*. Note, those with *μ* = 0, by assumption, cannot infect or get infected, and as such, for them, *e* = 0. Therefore, the choice *e* ∊ {0,1} is germane only for those who choose *μ* =1.

What are the costs of catching an infection? We posit that transitioning from AH to SI status causes a person to lose a fraction, *δ* ∊ (0,1), of her income presumably due to diminished productivity during convalescence. Neither the AI nor the AH incurs this loss.

### 2.2 Meetings

As explained in the introduction, all meetings are bilateral and can occur only between the AH and the AI people who have chosen *μ* =1. Such encounters are the only source of new infections. This is where two-sided prevention is key: the protection choices *e* ∊ {0,1} of each party matter in the possible transmission of the disease.

Let the superscripts *h* and *s* denote a AH and AI agent respectively. For future use, let

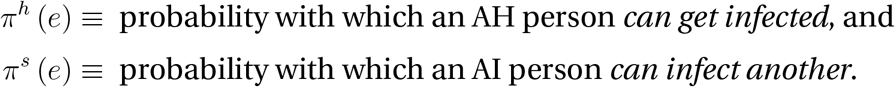

Notice, these probabilities are conditional on own protection choices. For example, *π^h^*(1) is the likelihood a self-protected, healthy agent gets infected from a random meeting; likewise, *π^s^* (0) is the probability with which an unprotected, infected person infects others. We assume {*π^h^*(1), *π^h^* (0), *π^s^* (1), *π^s^* (0)} are known from science and epidemiology, and accepted by all as common knowledge. By assumption,

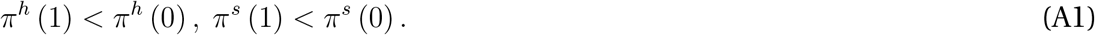

All else same, not engaging in protection increases one’s odds of both getting infected and of infecting others.

For future use, define

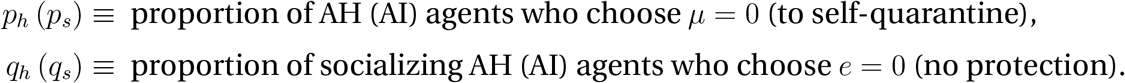

These probabilities will be derived endogenously from agent choices below. Also, notice *x*, *k, κ*, *λ*, *p_h_*, *p_s_*, *q_h_*, *q_s_*, *π^h^*(1), *π^h^*(0), *π^s^*(1) and *π^s^*(0) are the same for all *i*, in particular, the same across the rich and the poor. Constraining each of these to be independent of *w* imposes discipline on the exercise; it also allows us to focus on the connection between the distribution of *w* and infection rates.

For concreteness, the three types and their choices are as follows.

1. **AH**: Asymptomatic healthy, (1 − *α*)*h*
  a. self-quarantine (*p_h_*); *μ* = 0 Λ *e* = 0; cannot get infected.
  b. go out (1 − *p_h_*); *μ* = 1
    i. protection *e* = 1: fraction, 1 − *q_h_*; own infection probability, *π^h^* (1)
    ii. no protection *e* = 0: fraction, *q_h_*; own infection probability, *π^h^* (0)
2. **AI**: Asymptomatic, infected *αh*
  a. self-quarantine (*p_s_*); *μ* = 0 Λ *e* = 0. Cannot infect others.
  b. go out (1 − *p_s_*); *μ* = 1
    i. protection *e* = 1: fraction, 1 − *q_s_*; infect others with probability, *π^s^* (1)
    ii. no protection *e* = 0: fraction, *q_s_*; infect others with probability, *π^s^* (0)
3. **SI**: Symptomatic, infected (1 − *h*): *μ* = 0 Λ *e* = 0. Cannot further infect others.

Note, by assumption, the SI are removed from further analytical consideration. Also recall, the AH and the AI are unaware of their infection status but know *α*. Who are the people who are out and about? They comprise some of the AH, (1 − *p_h_*) (1 − *α*) *h*, and some of the AI, (1 − *p_s_*) *αh*. Only the former can get infected, and only the latter can infect. It follows, the fraction of infected people out and about, or the probability of meeting an infected agent, is

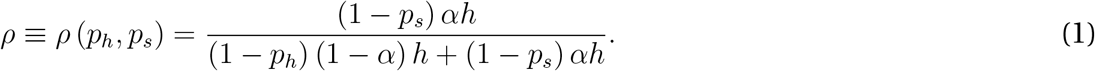

In a standard SIR model, where rational-choice considerations do not enter, the fraction of infected people would be *α* if the SI were under quarantine. Here, that fraction depends on both 1 − *p_h_* and 1 − *p_s_*, respectively, the proportions of AH (AI) agents who *choose* to go out.

### 2.3 Preferences and budget

Agents value consumption, *c*, using a utility function *u* (*c*). Agents’ consumption will depend on their infection status and the choices they make regarding *e* and *μ*. Overall, an agent’s utility function is

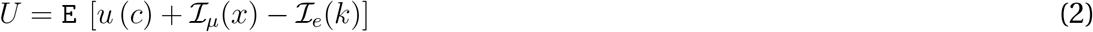

and his budget constraint

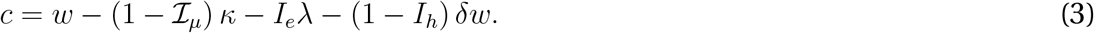

where 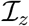 is an indicator function which takes on the value 1 when *z* =1 and 0 otherwise for *z* = *μ*, *e*, *h*. All decisions are based on *ex ante* expected utility where the expectation, E, is taken on the binary (infected or not) probability distribution. The agent maximizes (2) subject to (3), taking as given {*p_h_*, *p_s_*, *q_h_*, *q_s_*}.

## 3 Choices

Next, we figure out the choices made by agents on the dual extensive margins, going out and protection.

### 3.1 Unprotected socialization

Among the non-SI who go out, each agent can make two choices *e* = 0 (no protection) and *e* = 1 (protection). As mentioned above, the non-SI, i.e., AH and AI, unaware of their infection status, face the same *ex ante* decision problem.

First, consider a non-SI with *e* = 0. The agent has to take into account that she may be either AH or AI type, she can get infected only if she is the former and that her chance of getting infected depends on meeting an AI agent whose protection choices she does not know *ex ante*.

With probability *α*, this agent is AI: irrespective of whom she meets, her own health status is unchanged, and utility is *u*(*w*)+ *x*. With the remaining probability, (1 − *α*), this person is an AH, and now it matters whom she meets. With probability, (1 − *ρ*), she meets another AH, health status is unchanged and utility is *u*(*w*)+*x*. With the remaining probability, *ρ*, she meets an AI. Now her partner’s choice of *e* matters. If both parties have chosen *e* = 0, then she gets infected with probability *π^h^* (0) *π^s^* (0) with associated utility *u*((1 − *δ*)*w*) + *x*; her health status is unchanged with probability 1 − *π^h^* (0) *π^s^* (0). If the partner had, instead, chosen *e* =1, she gets infected with probability *π^h^* (0) *π^s^* (1) with utility *u*((1 − *δ*)*w*) + *x*, and her status is unchanged with the remaining probability, 1 − *π^h^* (0) *π^s^* (1). This is the power of two-sided prevention.

Recall, we defined the proportions of AI with (*μ*, *e*) = (1,0) and AI with (*μ*, *e*) = (1,1) as *q_s_* and 1 − *q_s_*, respectively. Since q_s_ is the probability of meeting an unprotected AI agent, an AH attaches the probability {*q_s_π^s^*(0) + (1 − *q_s_*)*π^s^*(1)} to a random AI person being capable of spreading infections. Then, the agent’s expected utility from going out, unprotected, conditional on her being AH and meeting an AI agent is

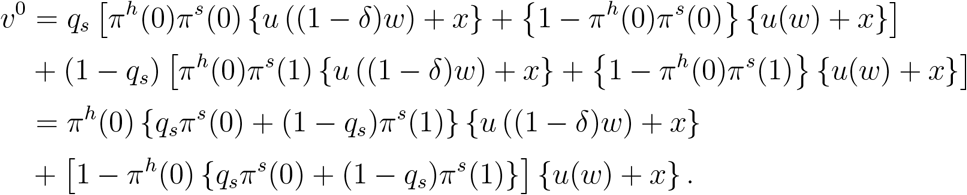

Hence, her expected utility from going out, unprotected, conditional on being AH is

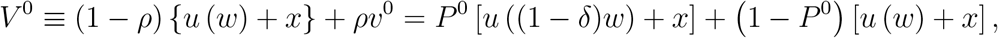

where

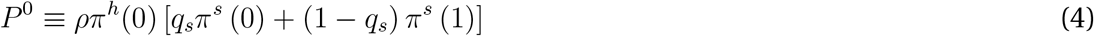

is the probability of getting infected for unprotected AH agents.

The probability that an asymptomatic person is infectious is *α*. Therefore, using the expressions above, the expected utility of a non-SI person (unknown infection status) who goes out, unprotected, is

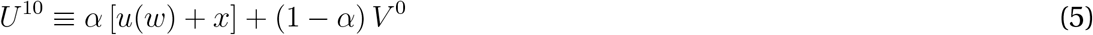

where we use the notation, *U^μe^*, to denote the expected utility from the choice of (*μ*, *e*) ∊ {0,1} × {0,1}.

### 3.2 Protected socialization

Analogously, for an AH agent, the expected utility of going out with protection is

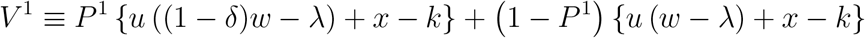

where

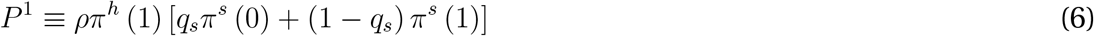

is the probability of getting infected of a protected AH agent. Notice, using (A1),

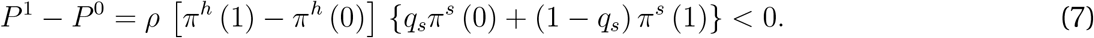

Intuitively, this is the reduction in the probability of contracting the disease achieved via self-protection while socializing. Therefore, a non-SI agent’s expected utility from protected socialization is

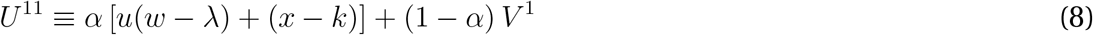

From (5) and (8), we conclude that, conditional on choosing to go out, a non-SI agent engages in protection iff *U*^11^ > *U*^10^.

### 3.3 To self-quarantine or not

It remains to understand, who among the non-SI agents goes out and who self-quarantines. If a non-SI agent is under self-quarantine, i.e., *e* = 0, his utility is

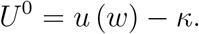

If he goes out, unprotected, his utility is *U*^10^. If he goes out, protected, his utility is *U*^11^. A non-SI agent chooses to self-quarantine if *U*^0^ > max {*U*^10^, *U*^11^} = *U*^1^.

We are now ready to compute the five masses of people that populate the economy:

i. AH who go out and protect,
ii. AH who go out, unprotected,
iii. AI who go out and protect,
iv. AI who go out, unprotected, and
v. those who self-quarantine.

It bears emphasis that membership in these categories depends on the *endogenous* probabilities, *p_h_*, *p_s_*, *q_s_*, *q_h_* as well as on *α* and *h*.

### 3.4 Protection decisions

Since protection is costly and does not bring any benefit to an agent under self-quarantine, we need only consider the protection decision for an agent who has chosen to socialize. Since AI and AH agents do not know their infection status, both *p_s_* = *p_h_* and *q_s_* = *q_h_* obtain which means

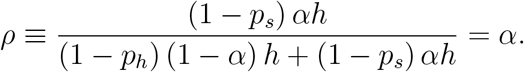

The utility difference, from protection versus not, for these agents, conditional on them having chosen to socialize, is:

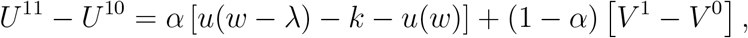

where

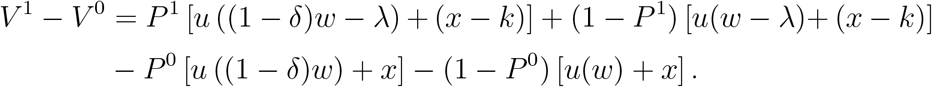

## 4 Closed form analytics

A road map of where we are headed is in order. We have established the margins on which the twin decisions of going out and prevention will hinge on. These decisions, as noted before, depend on the masses of the various non-SI pools which agent *i* takes as given. These masses, in turn, depend on the *endogenous* probabilities, *p_h_*, *p_s_*, *q_s_*, *q_h_*. At the individual level, as will be apparent below, agent *i*’s income will matter because of the goods costs associated with the choices. We will impose rational expectations on the equilibrium. This means, taking as parametric *p_h_*, *p_s_*, *q_s_*, *q_h_*, each agent *i* will make his selections, and ultimately, in the aggregate, those selections will validate the initial assumptions made by the agents. To make progress towards that, we make some simplifying assumption.

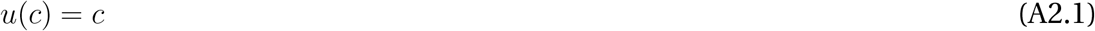

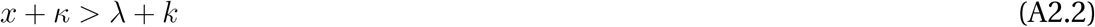

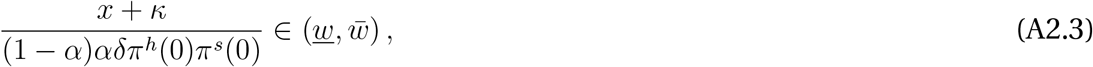

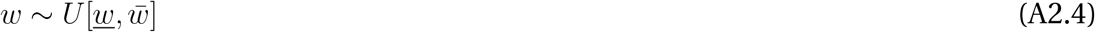

The advantage of (A2.1) is closed-form solutions, which cannot be obtained otherwise. By assuming the gains from socialization are sufficiently large, (A2.2) produces situations where some *i* will choose to both socialize and self-protect. The relevance of the other assumptions is addressed in due course.

### 4.1 Income cut-offs

For now, take as parametric *p_h_*, *p_s_*, *q_s_*, and *q_h_*. It follows for any agent *i*,

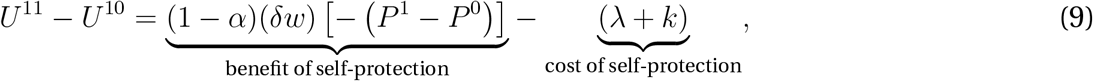

where *P*^1^ − *P*^0^ is given by (7). Because *P*^0^ > *P*^1^, the first term on the right-hand side of (9) is positive, and constitutes the expected benefit from self-protection. *δw* is the income (utility) loss avoided from protecting oneself, and *P*^0^ − *P*^1^ is the reduction in the probability of contracting the disease via self-protection while socializing, and (1 − *α*) is the probability the agent is AH. Because utility is linear, resource and disutility costs of protection are indistinguishable; only *λ* + *k* matters.

It follows from (9) that

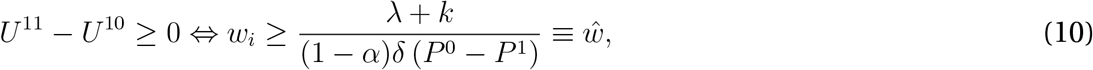

or, equivalently,

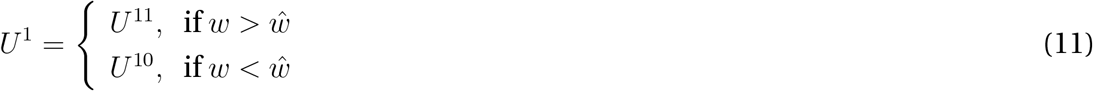

where *ŵ* is the same for all *i*. The larger the gains from protection and smaller the costs, the lower is the cutoff *ŵ*. Clearly the rich are more likely to invest in self-protection while socializing. This is because the income loss from falling sick from the disease is higher for richer agents while the cost of protection is independent of an agent’s income.

If an agent of any type is under self-quarantine, his utility, as noted earlier, is

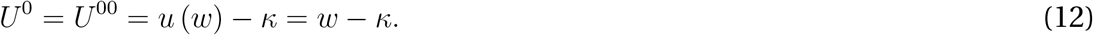

An AH or AI agent will socialize, with or without protection, as long as max{*U*^10^, *U*^11^} > *U*^0^ where, from (11), utility from socializing depends on *ŵ*. First, consider *w_i_* > *ŵ* which means *i* will protect himself should he choose to socialize. His expected utility gain from socializing under (A2.1) is

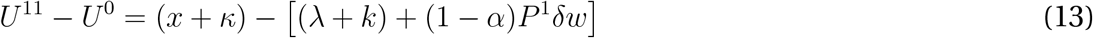

Here, the first term on the RHS is the net expected benefit of going out with protection: doing so avoids the income loss *κ* from self-quarantine and brings in a utility gain *x*. The second term is the effective cost of protection, *λ* + *k*, and the expected income loss from exposing oneself to infection. By (A2.2),

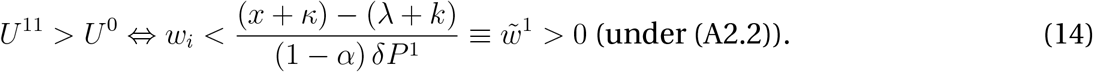

In other words, agents with 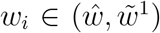 engage in protected socialization. For *w_i_*, < *ŵ*, on the other hand, going out without protection brings the net gain

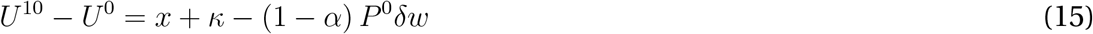

which is positive as long as

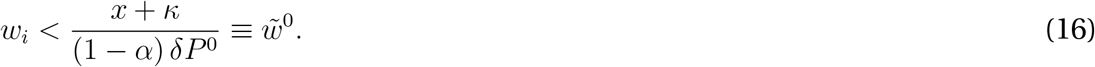

For agents with *w_i_*, < *ŵ*, unprotected socialization is optimal as long as 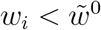.

A precise specification of prevention decisions requires the relative ranking of the three cutoffs, 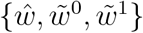. Observe that

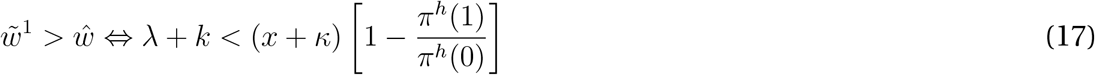

by (A1), while

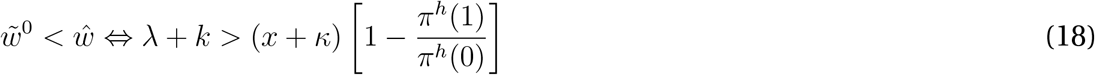

which also implies 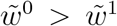. The last term within square brackets on the right-hand side of both inequalities is the proportional reduction in infection risk from protecting oneself.^9^

Prevention choices are completely characterized by the proposition below and illustrated in Figure 1.

**Figure 1:**
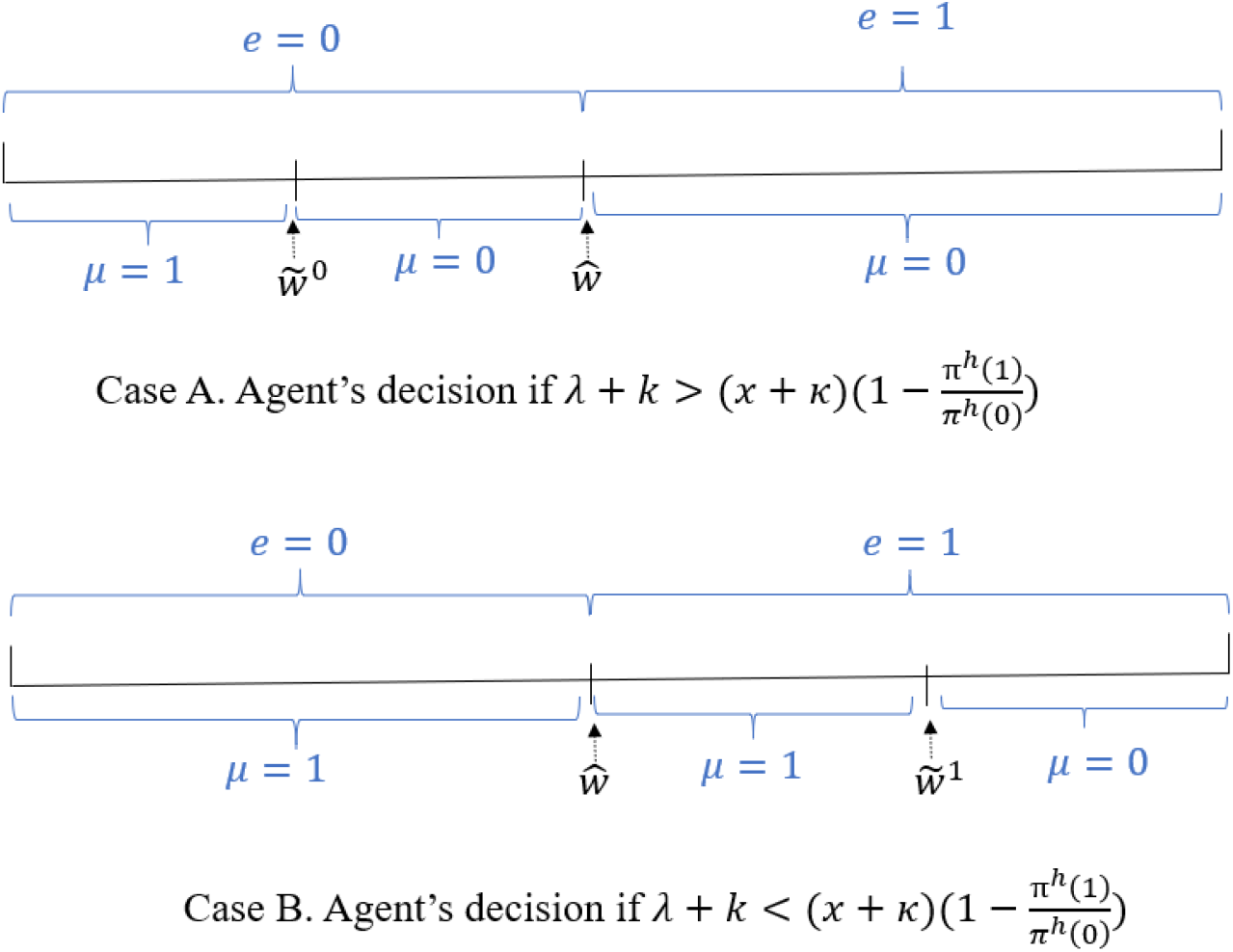
Prevention choices

#### Proposition 1

*Given the probabilities {ph,p_s_, q_h_, q_s_}, protection choices depend on how much protection reduces the risk of infection*.

A. *If λ* + *k* > (*x* + *κ*) [1 − *π^h^*(1)/*π^h^*(0)], 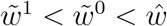,
  - *Agents with* 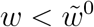 *choose* (*μ*, *e*) = (1,0): ***unprotected socialization***,
  - *Agents with* 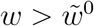 *choose* (*μ*, *e*) = (0,0): ***self quarantine***.
B. *If λ* + *k* < (*x* + *κ*) [l − *π^h^*(1)/*π^h^*(0)], 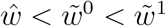,
  - *Agents with* 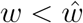 *choose* (*μ*, *e*) = (1, 0): ***unprotected socialization***
  - *Agents with* 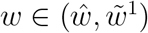 *choose* (*μ*, *e*) = (1, 1): ***protected socialization***
  - *Agents with* 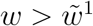 *choose* (*μ*, *e*) = (0, 0): ***self quarantine***.

## 5 Equilibrium

We now turn to the equilibrium determination of the probabilities {*p_h_*, *p_s_*, *q_h_*, *q_s_*}. This depends on the two cases described in Proposition 1. First note that, the (1 − *h*) SI proportion are assumed to be randomly drawn from the population. As they play no role in disease transmission in the model, all proportions below pertain to the non-SI, *h* fraction of the population.

In case A, all socializing is unprotected implying *q_h_* = *q_s_* = 1. It follows

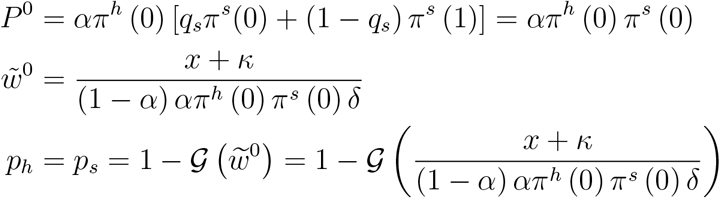

Assumption (A2.3) ensures *p_h_*, *p_s_* ∊ (0,1).

In case B, recall that a measure 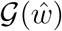 of agents choose to socialize without protection, while a measure 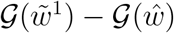 socialize with production. Therefore,

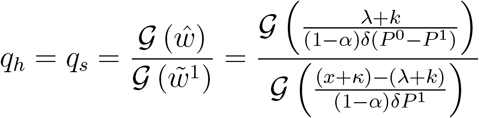

where 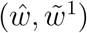 are functions of *q_s_* since *P*^0^ and *P*^1^ depend on *q_s_*. Also,

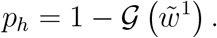

Here on, we make use of the uniform income-distribution assumption. Using, assumption (A2.4), we have

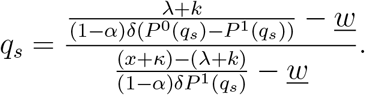

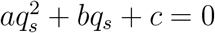 where

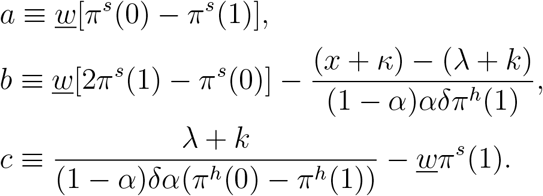

Depending on parameter values, the equilibrium *q_s_* is either zero or an interior (real) value 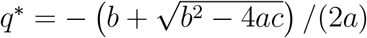 defined above.

The proposition below collects results for equilibrium (*p_h_*, *p_s_*, *q_h_*, *q_s_*) that are formally established in Appendix A.

### Proposition2

*Under* (A2.3) *and assumption* (A2.4)*, the equilibrium values* of (*p_h_*, *p_s_*, *q_h_*, *q_s_*) *are given by*

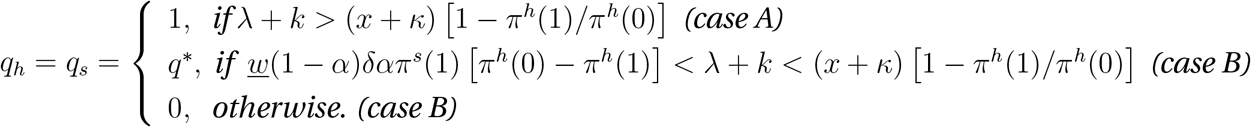

*and*

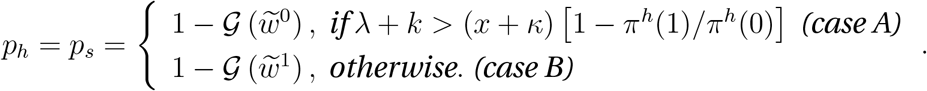

A side-note on multiplicity of equilibria is in order. Notice, the equilibrium values of (*p_h_*, *p_s_*, *q_h_*, *q_s_*), given a set of exogenous parameters, is unique. In Chang and Velasco’s (2020) model, multiple expectational equilibria in prevention can occur: if agents anticipate others not to engage in prevention, their expectation of high infection risk lowers their own incentives to protect themselves. Such an outcome is not possible here. This is easily appreciated by considering how much infection risk can be reduced by using protection during social interactions. Suppose agent i perceives the risk of infection to be high because he anticipates *q_s_* – the proportion of socializers who are unprotected – to be high. The gain to him from using protection is, *P*^0^ – *P*^1^ = *α* [*π^h^*(0) − *π^h^*(1)] [*q_s_π^s^* (0) + (1 − *q_s_*) *π^s^* (1)], *increasing* in *i*’s expectation of *q_s_*. It is this absence of strategic complementarity that prevents the possibility of multiple equilibria in prevention and prevalence.

Next, we conduct some comparative statics exercises. In our set up, these can be tricky because a marginal change in some of the shifters can alter, both, whether case A or B obtains, and the position of the income cutoffs 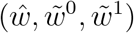. We start by asking, *ceteris paribus*, how do the various costs associated with self-protection affect prevention in equilibrium? Because the subjective utility costs *(k,x)* affect choices in a way similar to the resource costs (*λ, κ*), we consider only the latter. We appeal to (A2.4) to obtain analytical solutions for *(q,p_h_)* (further developed below) and illustrate the effects in Figure 2.

**Figure 2:**
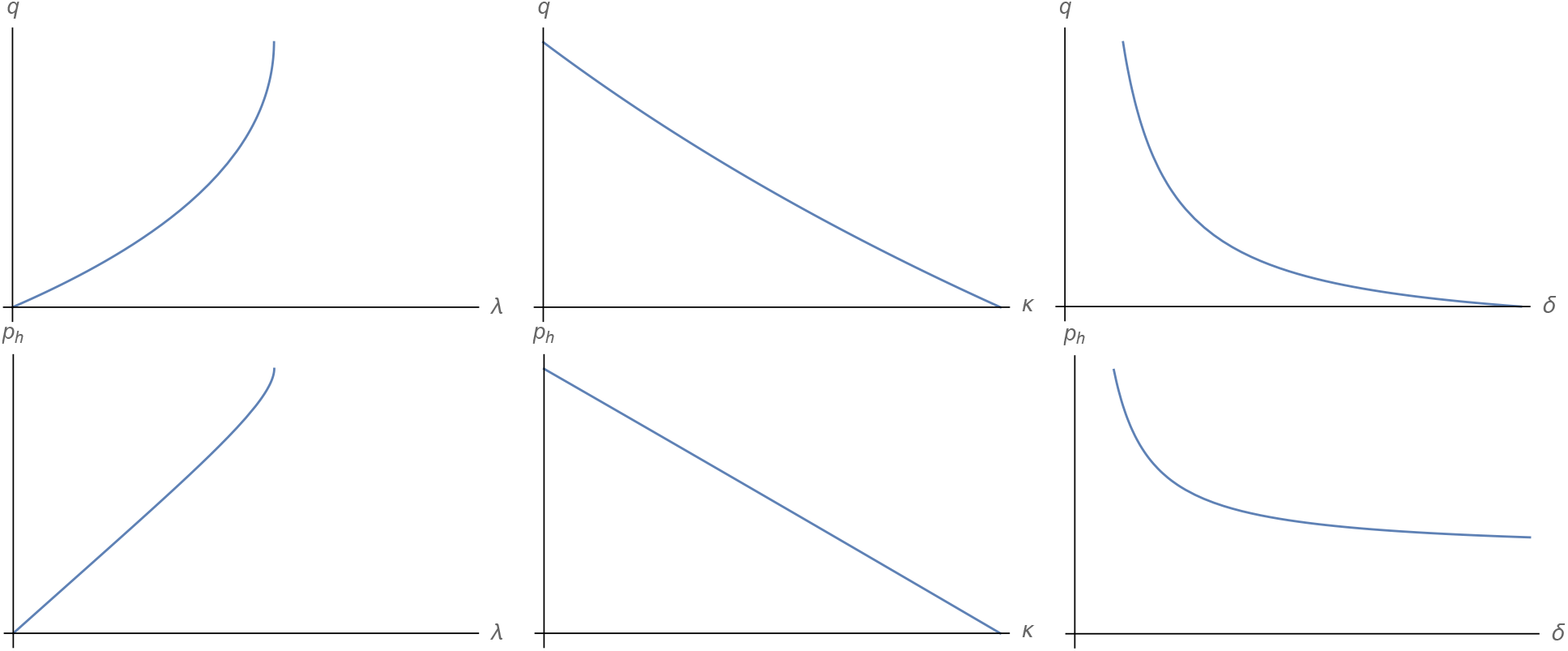
Effect of (*λ, κ*, *δ*) on equilibrium *q* (top row) and *p_h_* (bottom row)

Higher values of *λ* make protection-while-socializing costlier: in equilibrium fewer socializers opt to protect themselves (*q*) and, because that raises the risk of infection, a higher proportion of non-SI agents quarantine themselves (*p_h_*). The opposite is true for higher values of *κ*: fewer people self-quarantine, because it is costlier. This change occurs among relatively rich agents who are more likely to invest in protection; hence, more socializers protect themselves. These are the patterns we see above.

Also relevant is the cost of the disease itself, *δ* ∊ (0,1). Higher values of *δ* make non-prevention costlier. More and more socializers protect themselves as *δ* rises – *q* falls – while, in Fig 2, fewer of the non-SI agents socially isolate – *p_h_* falls too. The latter is surprising as both 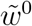 (case A) and 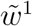 (case B) fall which ought to incentivize more agents to self-quarantine. The answer lies in the equilibrium effect of *q* on the risk of infections. As more socializers choose to use protection, the average probability of getting infected from socializing falls. This encourages some previously quarantined agents to go out with protection.^10^

## 6 Further Analysis

### 6.1 Behavior and infection transmission

In the standard SIR model, two variables are often used to measure the progression of the disease. The first is *R*_0_, the basic reproduction number. It is the average number of new infections generated by an infectious person in the absence of immunity and deliberate prevention steps. The other measure is *R*, the effective reproduction rate when some prevention methods are being used.

Disease transmission here differs from that in the SIR model along three margins. In the SIR model, it is implicitly assumed that everyone meets everyone else. Hence *S* susceptible people meet *I* infectious people, resulting in *S* × *I* opportunities for disease transmission, with an exogenous probability *β* of transmission at each meeting. This creates *βSI* new infections per *I* infectious person. In contrast, each agent in our model meets only one other person at random who may or may not be of a different type. Secondly, because agents can choose to self-quarantine themselves, not all such bilateral meetings between agents of different types happen. Thirdly, also part of prevention, the transmission rate of the disease is affected by the protection choices of a socializing susceptible agent and infective agent.

Our one-period model is not designed to track the dynamic evolution of the disease.^11^ Nevertheless, we can compute how many infections are generated by each infected agent. In the absence of self-quarantining or protection, there are *αhN* AI agents and (1 − *α*)*hN* AH ones. Hence, the average number of infections created by an infectious agent is

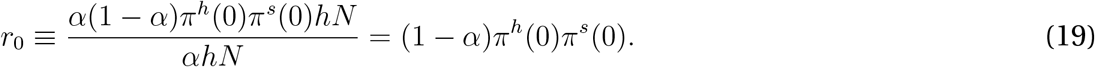

Notice, our *r*_0_ depends solely on the inherent contagiousness of the virus and broader demographic factors; deliberative human behavior plays no role.^12^ Note also *r*_0_ ≤ 1: in our setup with a single bilateral meeting, each infectious agent can meet at most one other person.

We can adjust this measure for behavior. Let

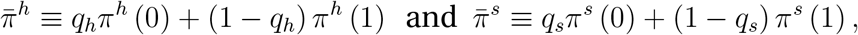

denote the average probabilities of getting infected and transmitting infections respectively. In equilibrium, a proportion 1 − *p_h_* of the AH and AI agents socialize. Some of them also use protection which we account for in the definitions of the average probabilities above. Therefore the number of infectious agents is *α*(1 − *p_h_*)*hN* and they infect 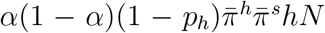 of the AH agents. In this case, the number of new infections created by each infectious agent is

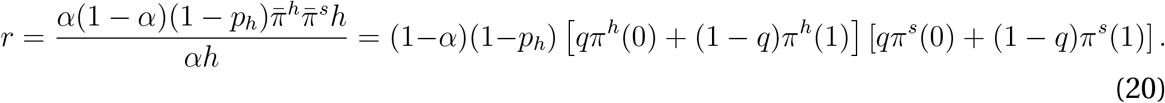

while that created by each *infective* agent, that is, a non-quarantined AI agent capable of transmitting the disease, is

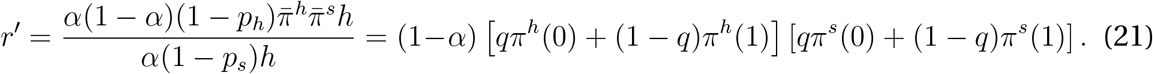

The difference between *r* and *r*′ lies in prevention methods. The latter is looking solely at how many infections are created in social settings. The former recognizes that self-quarantine is another tool to reduce spread and asks how many new infections are created per infectious person under both types of prevention.

Conditional on *q*, an increase in *π^h^*(0) obviously weakens prevention incentives and increases *r* and *r*′ while the opposite is true for an increase in *π^h^*(1). In equilibrium, q will respond to the change and may amplify or weaken these partial equilibrium responses. More interesting than these are the effects of protection (*k*, *λ*) and social isolation cost (*x*, *κ*) and disease cost *δ*. In Figure 3 we report how changes in these parameters affect equilibrium transmission (*r*, *r*′). The behavior of *r*′ follows directly from Figure 2 while the behavior of *r* is sensitive to parameter values as it is the interaction of the changes in *q* and *p_h_* in Figure 2.

**Figure 3:**
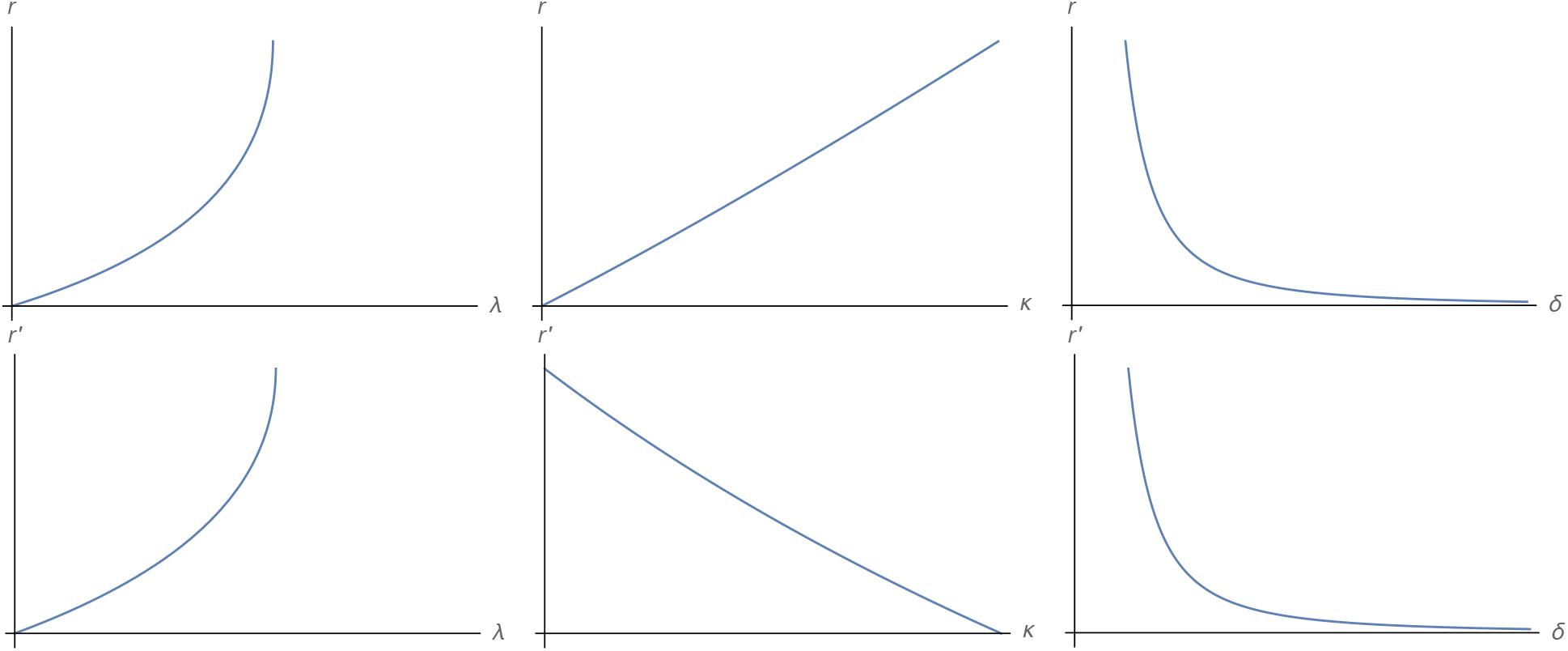
Effect of (*λ*, *κ*, *δ*) on equilibrium *r* (top row) and *r*′ (bottom row)

What is less apparent from the discussion so far is that the income distribution too affects choices and equilibrium disease transmission. We take that up below.

### 6.2 Effects of inequality

We study the effect of inequality on prevention behavior by focusing on the interior solution *q*^*^, that is when

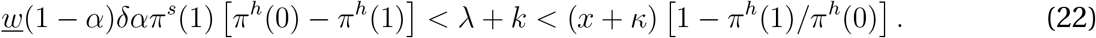

With slight abuse of notation, define 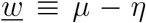 and 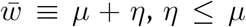, so that mean income is *μ* and variance *η*^2^/2. An increase in n constitutes a mean-preserving increase in inequality.

The (interior) equilibrium share of unprotected socializers, *q*^*^, now solves

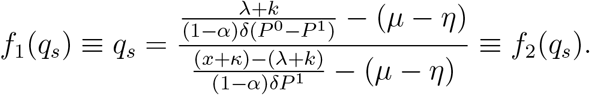

Straightforward differentiation shows that, under (22), ∂*f*_2_(*q_s_*)/∂*q_s_* < 0 and that ∂*f*_2_(*q_s_*)/∂*η* > 0. Hence, as shown in Figure 4, ∂*q*^*^/∂*η* > 0. A mean-preserving increase in inequality raises the proportion of socializing agents who are unprotected.

**Figure 4:**
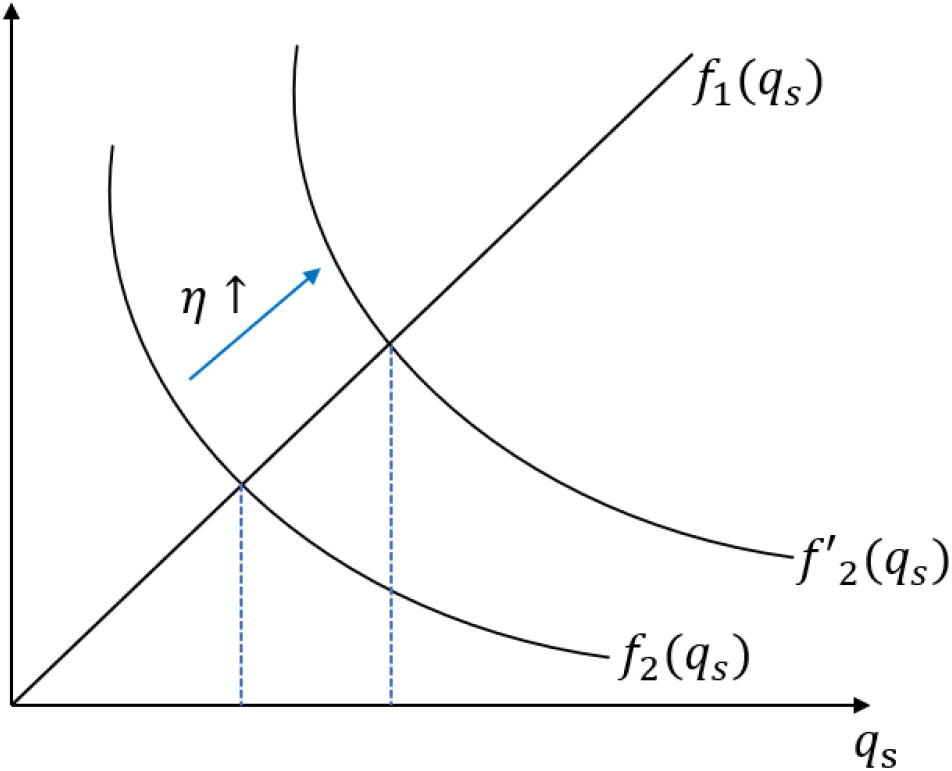
Higher inequality increases unprotected behavior

The equilibrium share of agents who self-quarantine, *p_h_*, is

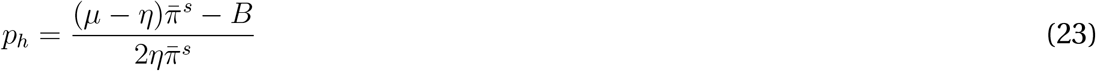

where *B* ≡ [*x* + *κ* − (*λ* + *k*)]/ [*α*(1 − *α*)*δπ^h^*(1)] and which may increase or decrease from an increase in *η*.

These results stated in Proposition 3 are formally established in Appendix B.

#### Proposition 3

*Under assumption* (A2.4)*, a mean preserving increase in the spread of the distribution of income increases equilibrium q_s_, the proportion of socializing agents who are unprotected, and may increase or decrease equilibrium p_h_, the proportion of agents who self-quarantine. While the overall effect on disease transmission is ambiguous, the risk of infections in social interactions is unambiguously higher*.

Of interest, too, is the effect of inequality on the propensity to transmit infections, (*r*, *r*′), as specified in equations (20) and (21). As both 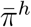 and 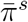 are increasing in *q_s_*, an increase in *η* increases *r* via *q_s_*. An increase in *p_h_*, on the other hand, lowers *r* but has no effect on *r*′. The net effect on *r* is ambiguous, though it stands to reason the first effect may dominate because it is amplified through the product 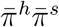.

Figure 5 shows the behavior of (*q_s_*, *p_h_*, *r*) as inequality (*η*) rises based on a set of parameter values. The main parameter that is varied across these figures to produce different outcomes is *x*, the subjective cost of social isolation. While *q_s_* always falls with inequality as shown in Proposition 3, the effects on *p_h_* and hence, *r* can differ.

**Figure 5:**
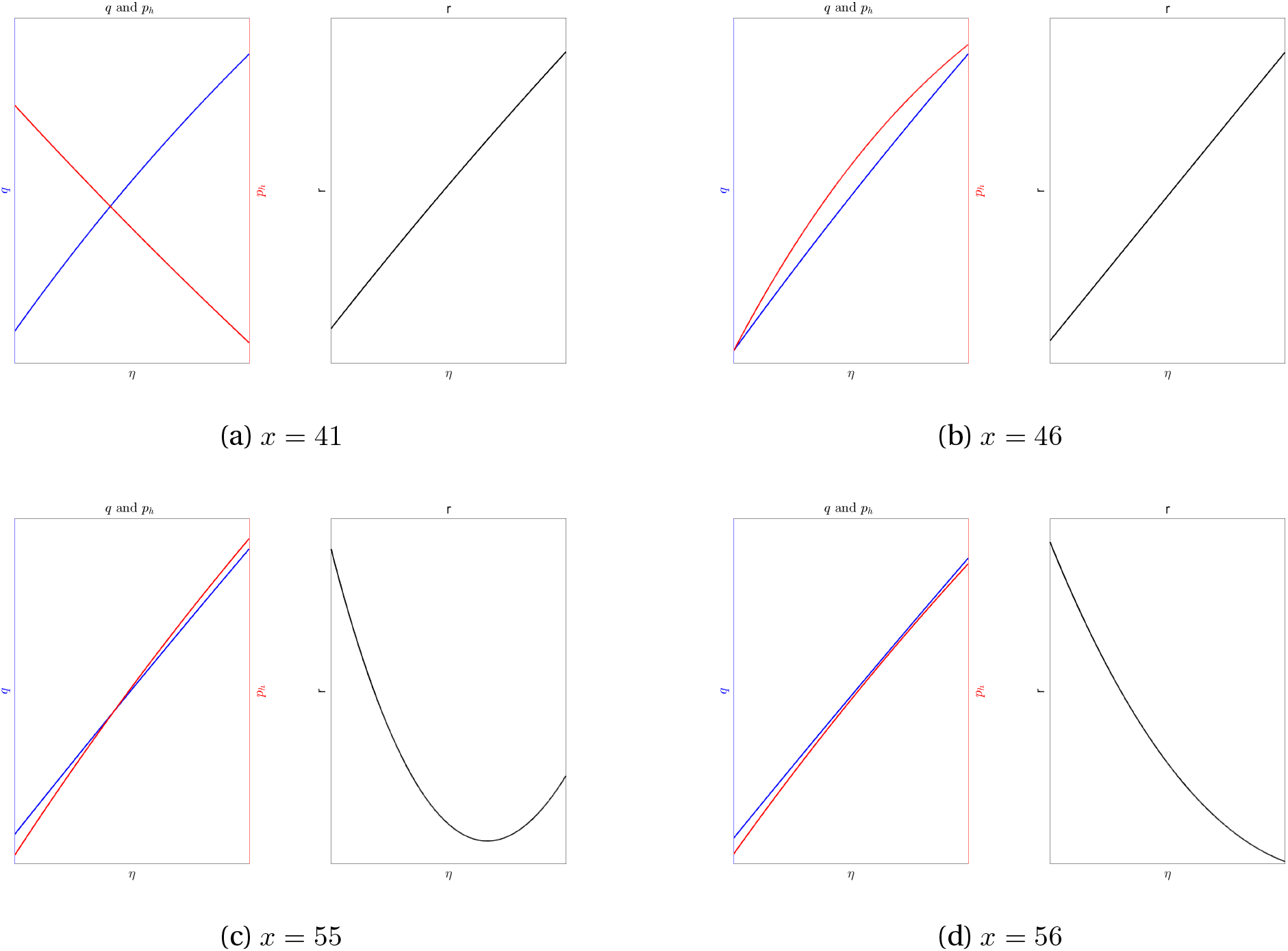
Effect of *η* on equilibrium (*q*, *p_h_*, *r*)

## 7 Discussion and Conclusion

Epidemiological models of the SIR type are extensively used to predict the evolution of contagion. Social scientists, especially in the context of human transmission, have long pointed to a significant lacuna in such models – the agents, therein, who catch or spread infections are essentially automatons. More gently, these models treat individual behavior as unresponsive and make no effort to incorporate people’s incentives to change behavior in the face of infection risks. This is a notable omission in the case of COVID because people, by now, are mindful that the primary transmission route is human-to-human and that vigilant usage of PPE can help.

This paper offers a minimalist, analytical model that allows for decision-making regarding socializing and prevention. It is specifically designed to study the positive (as opposed to normative) question: knowing about asymptomatic transmission, and not knowing about one’s own or the other party’s infectivity, would two people meet up to socialize, and if so, would they use PPE? These decisions are not trivial because a) selecting to go out in public, while utility-enhancing, appreciably raises one’s risk of contracting a very harmful infection, and b) not everyone likes to use PPE, at least not diligently. Not to mention, the income losses from self quarantining for long periods that some can ill-afford. We allow for agent heterogeneity in pre-COVID income and COVID status and analyze prevention choices and their effect on the transmission of infections. Our results highlight how the trade-off between cost and benefits of the two prevention choices, social isolation, and PPE, varies across the income distribution and depends on the behavior of other agents.

While the paper is silent on issues relating to policy design and impact, several key insights can be culled from the paper’s analysis. All else same, those who see the PPE as being too costly will eschew their use, and that raises the infection spread. But, in that case, cognizant of the infection risk, fewer people will go out in the first place, and that can reduce disease spread. The overall effect will depend on the masses of the groups, who protects and who steps out. What about a policy of mandatory mask use when out? At first glance, such a plan seems prudent. Upon deeper reflection, not necessarily so: knowing that others are using masks, which means infection risk conditional on going out is now lower, X may decide to venture out even if she had erstwhile decided against it in no policy world. That is, mandatory masking may increase the mass of socializers. Of course, mandatory lockdowns (no one can go out) will bring down the infection risk but are likely unsustainable, at least in a democracy, because it hurts the welfare of many (notably, the uninfected and the penurious median voter).

Our analytically tractable framework helps to identify the main trade-offs. But a more realistic model is required to understand how the trade-offs change over time, especially the policy trade-off inherent in tolerating a steep drop in income from social isolation versus the ability to contain the epidemic sooner than later. A quantitatively-useful model would marry our static framework into an otherwise standard, dynamic SIR model where the income loss from infection is explicitly modelled (e.g., Goenka *et al*.^13^ 2014). It is also possible to generalize the social isolation decision on the extensive margin by allowing agents to choose the number of social interactions, and to analyze the aggregate welfare consequences of various policy packages. We plan to take these issues up in future work.

Another avenue worth pursuing is disease and policy uncertainty. Being a new disease, our knowledge of COVID, it’s transmission routes, the efficacy of prevention choices, etc., is still preliminary. Conflicting data from vastly different countries and contradictory recommendations from policymakers add to that uncertainty. Reliable and consistent policy advice can, in this environment, be another tool to address the pandemic.

## Data Availability

no data was used

## A Appendix A: Proof of Proposition 2

Equilibrium *q_s_* solves the quadratic equation

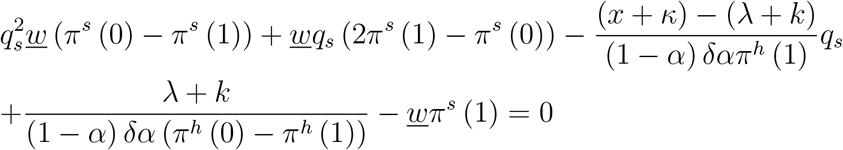

with two candidate solutions. Suppose 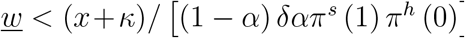. When 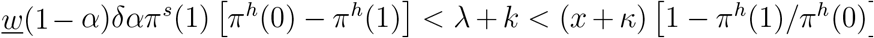, there exists and only exists one equilibrium in (0,1).

Define 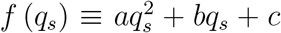, where a, b and c are defined in the main text. We have 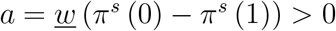 and

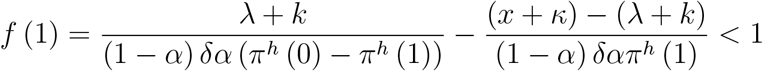

when *λ* + *k* < (*x* + *κ*) [1 − *π^h^*(1)/*π^h^*(0)]. Then a necessary and sufficient condition for a unique real solution to *f* (*q_s_*) = 0 in (0,1) is *f* (0) > 0. This is true as long as

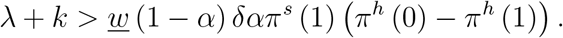

Under this condition, the equilibrium

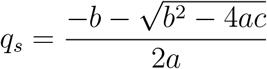

is an interior solution.

Notice the condition for *λ* + *k* < (*x* + *κ*) [1 − *π^h^*(1)/*π^h^*(0)] and 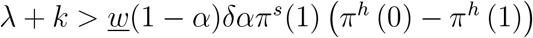 to hold requires that

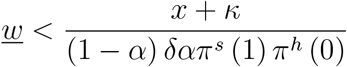

Assuming so, what if 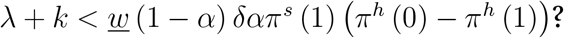

- Both *f* (0) and *f* (1) are negative and *f* (*q_s_*) ≠ 0 for *q_s_* ∊ (0,1). There is no interior equilibrium.
- If *q_s_* = 0, *P*^0^ = *απ^h^*(0)*π^s^*(1) and *P*^1^ = *απ^h^*(1)*π^s^*(1) and

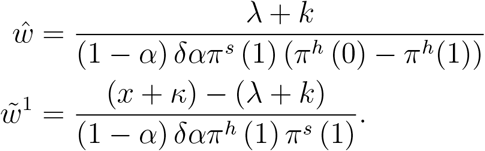 These imply 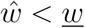 is satisfied. Therefore *q_s_* = 0 is an equilibrium.
- If *q_s_* = 1, *P*^0^ = *απ^h^*(0)*π^s^*(0) and *P*^1^ = *απ^h^*(1)*π^s^*(0) and

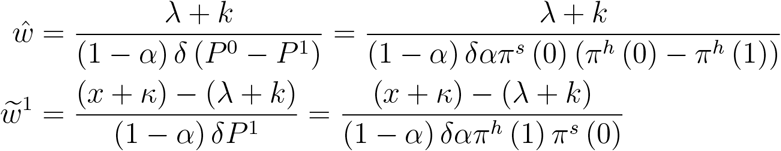 For *q_s_* = 1 to be an equilibrium, it must be that

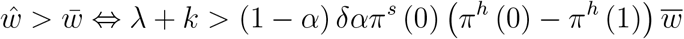 Since

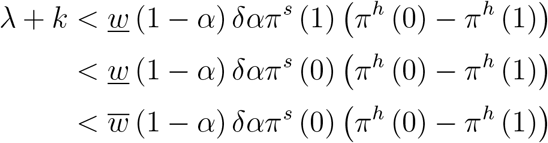

we have a contradiction and *q_s_* = 1 is not an equilibrium.

## B Appendix B: Proof of Proposition 3

We first show ∂*q*/∂*η* > 0. Rewrite

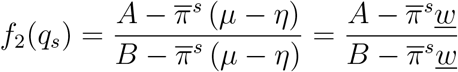

where, recall, *B* ≡ [*x* + *κ* − (*λ* + *k*)]/ [*α*(1 − *α*)*δπ^h^*(1)] and

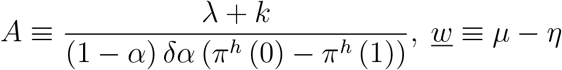

Then

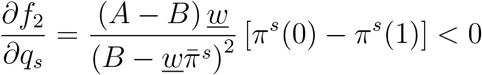

since *A* − *B* < 0 from (22). Similarly,

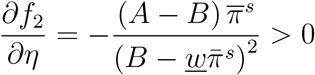

*q*^*^ solves *f*_1_ (*q_s_*) − *f*_2_ (*q_s_*, *η*) = 0. Hence

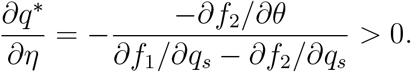

Next we show the sign of ∂*p_h_*/∂*η* is ambiguous. We defined 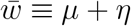. Then

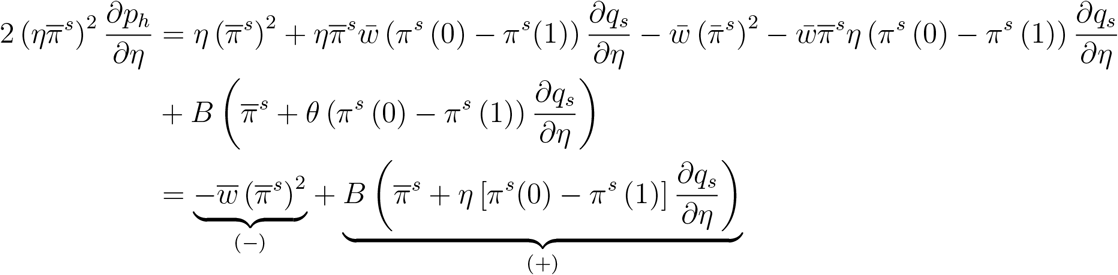

Hence ∂*p_h_*/∂*η* can be positive or negative. Beside other factors, 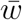 is likely important for the net effect.

## Footnotes

1 As empirical studies have indicated, people may be most infectious during the pre-symptomatic phase; see He *et al*. (2020) and Moghadas *et al*. (2020).

2 New contributions on these lines tailored for the COVID-19 pandemic include Acemoglu *et al*. (2020), Alvarez *et al*. (2020), Atkeson (2020), and Bethune and Korinek (2020).

3 Berger *et al*. (2020) also allow for incomplete information about infection status.

4 Farboodi *et al*. (2020) assume that utility depends on social activity while disease transmission depends on social interactions. That is, going out for a walk brings joy irrespective of whether any social interactions happen en route, but infections only occur when one group chooses to go out and happens to run into another group that has chosen to be out and about.

5 In reality, the AH mostly transition first to the pool of the AI; only later, some leave that pool and join the SI. For analytical convenience, we skip that initial step.

6 If the SI made prevention choices, they will always choose unprotected socialization based on selfinterest. Nothing substantive is, therefore, lost by focusing on the non-SI who have more interesting trade-offs to consider.

7 In reality, a person take a two-step decision: whether or not to venture out (extensive margin), and if so, howoften to engage with others (intensive margin). We lump all of the latter into one outing. Based on an online survey in the US, Canning *et al*. (2020) report that on average adults were in close contact with 1.9 non-household members the previous day. Interestingly they also find some evidence that having at least one COVID symptom increased the likelihood of going out.

8 A recent survey of American and British workers found that those in low-skilled occupations are typically able to performless of their duties from home (Abigail *et al*., 2020): while workers earning less than $40,000 per year can do less than 40% of their job from home, those earning more than $70,000 can do more than 60% (The Economist, 2020).

9 It can be sizeable. Chu *et al*. (2020), for example, estimate that masks alone cut down the risk of infection by 65%. If we include additional forms of protection such as physical distancing, face covering and hand-washing, even (18) is plausible despite the restriction placed by (A2.2).

10 It can be shown analytically that ∂*q_s_*/∂*δ* < 0, ∂*q_s_*/∂*λ* > 0 and ∂*q_s_*/∂*κ* < 0, while ∂*p_h_*/∂*λ* > 0 and ∂*p_h_*/∂*κ* < 0 with ∂*p_h_*/∂*δ* being ambiguous and depends on parameter values. For the parameter values used in Fig 2, the latter equilibrium effect dominates the direct effect of higher *δ* on *p_h_*.

11 The best way to do this would be to assume long-lived agents who are intertemporally forward-looking. Our agents are forward looking *within* the period because *ex ante* they attempt to evade infections by carefully weighing the static costs and benefits of their socializing and prevention decisions.

12 The “true” transmission rate is higher as the SI would all infect without protection if they could socialize.

13 Goenka et al. (2014) integrate a classic Ramsey model with the SIR framework. Their analysis works off the assumption that infected individuals are incapacitated and cannot work. This affects the labor force participation rate, and through that channel, affects all the major macroeconomic aggregates.

